# Transcatheter Aortic Valve Replacement (TAVR): A Comprehensive Descriptive Analysis

**DOI:** 10.1101/2025.07.10.25331325

**Authors:** Faria Nisar, Nicolas Mas D Alessandro, Alexander Estright, Samantha Pope, Daniel Schiller, Luis Tollinche, Maninder Singh

## Abstract

**Objectives:** TAVR is a minimally invasive alternative to SAVR, offering a key treatment for high-risk patients with severe aortic stenosis, including those with advanced age or comorbidities. Its popularity has led to its use in lower volume institutions, where outcomes are comparable to higher volume centers. This study analyzes procedural volume and outcomes at our institution, compares them with the PARTNER III trial, and evaluates complications and patient selection criteria.

**Design:** This was a retrospective descriptive study.

**Setting:** This study was performed at a public, multi-institutional healthcare system featuring an acute care hospital with a Level I Adult Trauma and Burn Center.

**Participants:** We obtained retrospective data on 37 patients who underwent TAVR in our hospital system in 2022. Data was collected on their outcomes, including mortality, readmission rates, and cardiovascular and cerebrovascular complications at 30 days and 1 year.

**Interventions:** No interventions were performed on patients as this was a retrospective study.

**Measurements and Main Results:** Patient outcomes were compared with PARTNER III trial results. There was no 30-day mortality (versus 1% in the PARTNER III trial), and a 1-year mortality rate of 2.7% as compared to 8.5% in the PARTNER III trial. At 30 days, 0% of patients were readmitted for valve-related issues.

**Conclusions:** This study demonstrates that lower volume healthcare institutions can achieve outcomes comparable/similar to higher volume institutions in specialized procedures like TAVR, challenging the assumption that superior outcomes are tied to institutional size. Our findings highlight the importance of quality, teamwork, and individualized care in achieving excellent clinical outcomes, regardless of size.

## Introduction

Transcatheter Aortic Valve Replacement (TAVR) has revolutionized the management of severe aortic stenosis, offering a less invasive alternative to surgical aortic valve replacement (SAVR) for high-risk patients [1], [2]. The evolution of TAVR from a novel procedure to a standard of care reflects its remarkable clinical impact and continuous advancements in technology and techniques.

Simultaneously, there has been a significant rise in the volume of procedures conducted and the number of facilities performing these procedures in both North America and globally[3]. This upward trajectory is expected to persist, given the projected increase in the elderly population with aortic valve disease, along with the expanding use of this technology in patients with lower surgical risk[3], [4].

A phenomenon known as the learning curve, characterized by enhanced outcomes with increasing procedural experience, has been observed across various cardiac and noncardiac procedures. Furthermore, maintaining a minimum annual procedural volume has been linked to better clinical outcomes [5].

The necessity for further development and consideration of this knowledge arises from research indicating a higher risk of adverse effects in settings where a specific medical procedure is not widely practiced and fails to meet the minimum required threshold. This risk can be mitigated by modifying the risk profile of patients selected [6], [7], [8], [9].

The objective of this study is to conduct a comprehensive analysis of our institution’s procedural volume, and outcomes over a 12-month period, and to compare them with those of other institutions. Furthermore, we aim to evaluate the associated complications with our primary outcome being mortality rate at 30 days and 1 year.

## Methods

In this retrospective descriptive analysis, we made a comparison between the TAVR outcomes of a lower volume institute like MH and major institutions. For this reason, we compared TAVR outcomes with “The PARTNER III Trial,” which is a multi-centered clinical trial involving seventy-one prominent institutions. The primary outcome to compare was “30-day and 1-year mortality” and secondary outcomes include patients’ length of hospital stay, conversion to general anesthesia, and post-operative complications (hematoma, bleeding, infection). We collected retrospective data of 37 patients from their electronic medical records who received TAVR procedures from MH from January 2022 to December 2022, representing all TAVR procedures performed at the institution during that year. Then, we analyzed various outcomes, including mortality, readmission rates, cardiovascular complications, and cerebrovascular complications for 30 days and 1 year using the Chi-Square Test and Fisher’s Exact Test.

All TAVR procedures were performed under monitored anesthesia care (MAC) or general anesthesia. After standard ASA monitoring and placement of an arterial line, sedation was initiated with a propofol infusion at 150-200 mcg/kg/min and dexmedetomidine at 0.5 mcg/kg/min, titrated to maintain an adequate sedation score. Fentanyl boluses were administered as needed for patient discomfort or hemodynamic responses. Sedative infusions were weaned upon completion of valve deployment, and patients were transferred to the recovery unit once they met criteria.

## Results

Between January and December 2022, 37 patients received the TAVR procedure in the MetroHealth System. The clinical outcomes at 30 days and 1 year were compared to the PARTNER III trial. During the initial 30 days following the TAVR procedure, it was observed that there have been no reported deaths at all in MH whereas there was a 1% mortality rate for patients involved in the PARTNER III trial with a p-value of >0.9. (Table 1) Moving forward to one-year outcomes, low mortality persisted in MH accounting for 2.7% while assessing understanding of the death rate observed in PARTNER III with 8.5% with a p-value of 0.3 (Table 1).

**Table 1.**

| **Characteristic** | **30 days** |  |  | **1 year** |  |  |
| --- | --- | --- | --- | --- | --- | --- |
|  | **MH,**<br>**N =**<br>**37<sup>1</sup>** | **PARTNER**<br>**III, N =**<br>**496<sup>1</sup>** | **p-**<br>**value<sup>2</sup>** | **MH,**<br>**N =**<br>**37<sup>1</sup>** | **PARTNER**<br>**III, N =**<br>**496<sup>1</sup>** | **p-**<br>**value<sup>2</sup>** |
| **Death from**<br>**any cause** | 0<br>(0%) | 5 (1.0%) | >0.9 | 1<br>(2.7%) | 42 (8.5%) | 0.3 |
| <sup>1</sup> n (%) |  |  |  |  |  |  |
| <sup>2</sup> Fisher’s exact test |  |  |  |  |  |  |

The patients’ length of stay had an average of 2.53 days (SD=2.45), a median of 2.00 days (Q1=1.25, Q3=2.00), and ranged from 1 to 11 days. Only 6 patients (16%) had a non-routine discharge after 3 days. The TAVR procedure duration averaged 83 minutes (SD=15), a median of 80 minutes (Q1=72, Q3=93), and a range of 52 minutes to 112 minutes. The length of time in the operating room had an average of 140 minutes (SD=30), a median of 135 minutes (Q1=125, Q3=147), and a range of 94 minutes to 273 minutes. None of the 37 cases were converted to general anesthesia and none had active bleeding. At the 30-day mark, not even a single patient was readmitted for valve-related reasons. These results can be seen in Table 2.

**Table 2.**

| Characteristic | N = 37<sup>I</sup> |
| --- | --- |
| **Length of Stay (days)** |  |
| Mean (SD) | 2.53 (2.45) |
| Median (25%,75%) | 2.00<br>(1.25,2.00) |
| Range | 1.00,<br>11.00 |
| **Non-routine discharge 3 days?** |  |
| N | 31 (84%) |
| Y | 6 (16%) |
| **Procedure Duration (min)** |  |
| Mean (SD) | 83 (15) |
| Median (25%,75%) | 80 (72,93) |
| Range | 52, 112 |
| **Converted to GA?** |  |
| N | 37 (100%) |
| **Active Bleed?** |  |
| N | 37 (100%) |
| **Readmission within 30 days (valve related)?** |  |
| N | 37 (100%) |
| Y | 0 (0%) |
| <sup>I</sup> n (%) |  |

## Discussion

In our evaluation of 37 cases undergoing transcatheter aortic valve replacement (TAVR) at MH, we noted significant results regarding procedural outcomes and patient recovery. Our findings revealed a hospital admission duration of 2.53 days. This is a shorter recovery time when compared to invasive cardiac surgery procedures. Shorter hospital stays have been linked to higher patient satisfaction and reduced healthcare costs. Notably, all procedures were completed without converting to general anesthesia. This highlights our institution’s proficiency in utilizing local anesthesia and mild-moderate sedation effectively during TAVR procedures.

These techniques have proven to be effective for patient comfort, recovery, and safety. Additionally, the absence of active bleeding during procedures in all cases underscores our meticulous procedural techniques and hemostatic control measures. Regarding early readmissions within 30 days for valve-related issues, none of the patients required readmission, suggesting effective post-procedural care and monitoring strategies. In addition, there were no deaths reported at one year which is a significant difference when compared to the PARTNER III trial. These findings suggest that our institution’s approach to patient-centric care, which involves close communication with the patient, surgical team, and anesthesia team, effectively mitigates risks associated with early readmission. Overall, our data highlight our institute’s effective management of TAVR procedures.

Despite its lower volume, MH produces statistically significant decreased mortality rates than higher volume institutions in the PARTNER III trial. This highlights that lesser volume institutions can provide high-quality care for specialized procedures, like TAVR. MH outperformed the PARTNER III trial overall in TAVR, possibly due to factors such as personalized patient care, dedicated multidisciplinary teams, and a focused approach to perfecting procedures on a smaller scale. These findings challenge the assumption that superior healthcare outcomes are solely linked to the size and resources of larger medical facilities, instead suggesting that successful clinical outcomes are driven by factors such as team cohesiveness, procedural experience, and individualized patient management. This research suggests a need for healthcare delivery systems to reevaluate their assumptions about an institution’s size relating to its quality of care, emphasizing quality metrics, patient satisfaction, and clinical outcomes rather than institutional size.

Further research is essential to understand what contributes to the success of MH in TAVR procedures, enabling us to improve patient outcomes and procedural efficiency. We are dedicated to refining our cardiovascular intervention strategies to enhance patient care through best practices. Further understanding on team experience, patient selection criteria, and perioperative care protocols among other variables would provide a better insight into clinical excellence drivers as well as improve cardiovascular practices. Moreover, longitudinal studies exploring the sustainability of outcomes during extended follow-up periods will further expound on the long-term values and challenges of TAVR use within different health systems.

Additionally, our findings address broader issues of healthcare quality, patient-centered care, and the ability of lesser volume institutions to manage complex interventions. Understanding these variations will help providers and policymakers develop tailored approaches to improve outcomes across different healthcare settings. In this way, a collaborative learning process is implemented, enabling people from different healthcare systems to share experiences on improving clinical outcomes while remaining patient-centered.

In summary, there are two key messages we can take away from our study. First, we note that tailored patient care and interdisciplinary teamwork must be encouraged in MH to achieve better TAVR results. Also, the PARTNER III trial was compared to how different healthcare settings led to a variation in outcomes. While larger trials give broad data, they may miss out on the subtle advantages possible in lesser volume, specialized environments. These subtleties should be understood to advance evidence-based practices and refine care protocols for various populations with diverse needs.

## Conclusion

Our study highlights the potential of lesser volume healthcare institutes like MH to excel in specialized cardiovascular procedures, challenging traditional notions of healthcare quality based on institutional size alone. We advocate for recognizing and supporting healthcare excellence irrespective of institutional size, emphasizing quality over quantity in healthcare delivery.

## Data Availability

Data for this project was procured the department and can be made available upon request.

